# Estimating the health impacts of sugar-sweetened beverage tax for informing policy decisions about the obesity burden in Vietnam

**DOI:** 10.1101/2022.09.08.22279712

**Authors:** Duyen Thuy Nguyen, Minh Van Hoang, Son Dao, Phuong Hong Do, Quang Dinh Nguyen, Jo Jewell, Ben Amies-Cull, Maharajan Muthu, Ly-Na Hoang, Thu Thi Le, An Thi Nguyen, Bao Quoc Tran, Ciaran O’Neill

## Abstract

**Background:** Considered a “best buy” intervention to cope with the obesity burden, a tax on sugar-sweetened beverages (SSBs) has been adopted in more than 40 countries. In Vietnam, a tax on SSBs has been proposed several times (most recently in 2017). This study aimed to estimate the health impacts of different SSBs tax plans currently under discussion to provide an evidence base to inform decision-making about a SSBs tax policy in Vietnam.

**Method:** Five tax scenarios were modelled, representing three levels of retail price increase: 5%, 11% and 19-20%. Scenarios of the highest price increase were assessed across three different tax designs: ad valorem, volume-based specific tax & sugar based specific tax. In each case we modelled SSBs consumption in each tax scenario; how this reduction in consumption translates to a reduction in total energy intake and how this relationship in turn translates to an average change in body weight and obesity status among adults by applying the calorie-to weight conversion factor. Changes in diabetes type 2 diabetes burden were then calculated based on the change in average body mass index of the modelled cohort. A Monte Carlo simulation approach was applied on the conversion factor of weight change and diabetes risk reduction for the sensitivity analysis.

**Results:** While the impact of a 5% price increase arising from a tax was relatively small, increasing SSBs’ price up to 20% appeared to impact substantially on overweight and obesity rates (reduction of 12.7% and 12.4% respectively) saving 27 million USD for direct medical cost. The greatest reduction in rates was observed for overweight (23≤BMI<25) and obesity grade 1 (25≤BMI<30). The decline in overweight and obesity rates was slightly higher for women than men. Differences were evident across all three tax designs with a specific tax based on sugar density achieving greatest effects.

## BACKGROUND

In 2016 it was estimated that globally almost half of adults and one fifth of children aged 5-19 were overweight or obese (1, 2). By 2030, it is estimated that over 1 billion people globally will be overweight or obese. (3). Low and middle-income countries observed the most rapid increase in obesity status in recent years (3). Therefore, it is critical for everyone, especially children, to access affordable and appealing healthy diets including avoiding or limiting health-harming drinks. There is strong, consistent evidence linking sugar-sweetened beverage (SSBs) consumption to weight gain and increased risk of overweight and obesity in children, adolescents, and adults (2, 4). There is evidence, too, showing that SSBs are the main source of added sugar in diets (4). It has been estimated that for every 100 million cases of type 2 diabetes predicted to occur in the US, about 8.7 million cases can be attributed to consumption of SSBs (5). In the UK, SSB consumption is estimated to be responsible for about 3.6 million cases out of every 100 million cases of type 2 diabetes that occurs (5).

Increasing taxes on SSBs has been strongly encouraged to reduce the SSB consumption as part of a comprehensive, evidence-based approach to improving diets and reducing the burden of diet-related NCDs (4, 6). So far, over 40 jurisdictions have levied a tax on SSBs in order to increase the price and reduce the consumption of this kind of beverage (4). According to the World Health Organization (WHO), SSBs are defined as all types of beverages containing free sugars and these include carbonated or non-carbonated soft drinks, fruit/vegetable juices and drinks, liquid and powder concentrates, flavoured water, energy and sports drinks, ready-to-drink (RTD) tea, RTD coffee, and flavoured milk drinks. Free sugars refer to monosaccharides (such as glucose, fructose) and disaccharides (such as sucrose or table sugar) added to foods and drinks by the manufacturer, cook or consumer, and sugars naturally present in honey, syrups, fruit juices and fruit juice concentrates (4, 7).

In Vietnam, while the rising trend of NCD burden has been observed in the last decade, Vietnam has shown limited progress towards achieving the diet-related non-communicable disease (NCD) targets for 2020 (8). A rapid increase in overweight and obesity status was observed among both children and adults. The prevalence of overweight and obesity almost doubled in adults (from 10.9% in 2002 to 18.3% in 2016) (9) and rose by 7 times in youths aged from 5-19 (from 2.6% in 2002 to 19% in 2020) (10). Over the same period, the consumption of SSBs has increased considerably. In the period 2002 - 2016, the consumption of carbonated drinks increased 3 times, juices increased 10 times, sport and energy drink increased 9 times and sweetened ready-to-drink tea increased 6 times (11).

To date, limited efforts have been made to limit trends in overweight and obesity. SSBs were considered for inclusion in the list of goods subjected to a special consumption tax (or excise tax) under Vietnam’s Law on Excise Tax in 2008 but it was not materialized. In 2017, the Ministry of Finance again proposed an ad-valorem tax at 10% applied on SSBs (12). In this draft law, a definition of SSB products was proposed to cover carbonated soft drink, non-carbonate soft drink, energy drink, sport drink, ready-to-drink tea/ coffee, except 100% juices, milk and milk-based drinks. This time it was again not adopted (12). To support decision-making about the development of SSBs tax policy, this study aimed to estimate the potential impacts on overweight and obesity and their main health effects of different SSBs tax plans under discussion in Vietnam.

## METHODOLOGY

### Model structure

**Error! Reference source not found**. below presents a theoretical framework for modelling the impact of SSBs taxation on the consumption of SSBs and the prevalence of obesity and overweight and a major related NCDs. The model structure was drawn based on the causal pathway of taxation policies on SSBs articulated in several studies in Thailand, South Africa, US, UK, Ireland, Mexico, Australia, Indonesia, Zambia (13-19).

As the study employed a multiple stage modelling approach, there are two time-horizons applied, starting with the 3-year horizon for weight change projection (short-term impact) and a lifetime horizon for the prediction of diabetes mellitus prevalence (long-term impact). For the first projected outcome regarding weight change and obesity status, the time horizon of our model was set at 3 years from the point of SSBs tax implementation based on the average time to obtain expected weight change (20).

### Model scenarios

In Vietnam, currently the excise tax is imposed on the ex-factory before-tax price (or factory price). The government tax strategy until 2030, No508/QÐ-TTg 2022, indicated that an excise specific tax component (as fixed amount of tax placed on a product quantity such as volume or sugar content) could be considered for some products so as to have a mixed ad valorem/specific tax system. Table 1 below illustrates five SSBs tax scenarios selected for modelling with average increase in retail price. The average price increase was estimated assuming that the amount of tax increase will be 100% passed on to the retail price. The five scenarios were (1) ad valorem tax of 10% on SSBs’ factory price; (2) specific tax at 3,500d (0.15 USD) per litre of SSB; (3) specific tax of 60d per 1 gram of sugar per 100 ml; (4) specific tax at 7,000d (0.30 USD) per litre of SSB and (5) ad valorem tax of 40%. These scenarios were designed to compare three levels of price increase 5% (scenario 1) vs 11% (scenario 2) vs 19-20% (scenario 3, 4, 5). Also, the three highest scenarios (scenario 3 to 5) allow a comparison of three designs of excise tax: ad valorem tax; volume-based specific tax; and sugar-based specific tax which give the same level of price increase at 19-20%. Specifically, scenario 1 was established based on the nearest proposed tax plan by Vietnam Ministry of Finance. Scenario 2 was designed as an in-between scenario for comparing the impacts of the proposed plan (scenario 1) and the desired plans (scenario 3, 4 and 5). Scenario 3, 4 and 5 was designed to meet the goal of increasing retail price by an average of 20% for an effective reduction in consumption as suggested by WHO (21).

**Table 1:** Selected tax scenarios for modelling.

| No | Tax type | Tax design | Interpretation | Tax rate | Price increase |
| --- | --- | --- | --- | --- | --- |
| 1 | Ad valorem tax |  | Tax as % of factory price | 10% | 5% |
| 2 | Specific tax | Volumetric | Fixed amount of tax imposed on product's volume | 3,500d/ litre | 11% |
| 3 | Specific tax | Sugar content | Fixed amount of tax imposed on grams of sugar contained | 60d/ gram/ litre | 19% |
| 4 | Specific tax | Volumetric | Fixed amount of tax imposed on product's volume | 7,000d/ litre | 20% |
| 5 | Ad valorem tax |  | Tax as % of factory price | 40% | 20% |

### Simulation techniques

#### Step 1 - Change in SSBs consumption

This step was designed to cover two transitions: (i) tax to price and (ii) price to consumption. In combination, the impact of tax policy placed on SSBs’ retail price is estimated, giving a prediction of reduction in SSBs consumption after imposing SSBs tax.

##### Step 1a: The transition from tax to retail price

This study used the findings from the TaxSim simulation model on sugar-sweetened beverages carried out by WHO & Health Bridge Canada in Vietnam (22) to obtain the average price increase in each tax plan. Data on the SSBs consumption and retail price was extracted from the Soft Drinks Volume and Value databases in 2020 from Global Data. This study estimated the SSBs consumption in four groups: (1) carbonates, (2) juice included liquid and solid state of juice, nectars, squash/syrups, still drinks, and fruit powders, (3) RTD tea and coffee, (4) sport and energy drinks. These figures included both regular and low-calorie product lines. The baseline consumption was collected for SSB products in four categories as above, measured in litres per day per person. The average retail price increase was estimated assuming that the pass-on rate from tax increase to factory price is 100% which equivalents to the pass-on rate of about 50% from tax increase to retail price.

##### Step 1b: The transition from price to consumption

The model employed the own-price elasticity of SSBs from Linh Luong et al 2020 which estimated price elasticities using Vietnam Household Living Standard Survey 2016 (23). The own-price elasticity of SSBs was estimated at -1.14 indicating that if the price of a SSB increased by 1%, each household would decrease their consumption of SSBs by 1.14% (23). This value was considered comparable with findings from a meta-analysis in low-and-middle-income countries and previous studies in Thailand or South Africa (15, 17, 24, 25). It is important to note that the price elasticity estimated for soft drink in Linh Luong et al 2020 (23) included both fresh water and other sugary drinks. The price elasticity was assumed to be the same for the whole population and assumed to be constant throughout the modelled time horizon.

#### Step 2 – Change in energy intake

We used average sugar content in each SSBs category multiplied by the average change in SSB consumption to estimate the change in the amount of sugar consumed from SSBs. The energy content in kilojoules (kJ) was calculated based on grams of sugar content. Then, the changes in caloric intake from SSBs were summed to give the net change in energy intake. To reduce model complexity, we assumed that no significant changes in other sources of energy intake happened.

The sugar content in carbonates, juices, RTD coffee and tea, and sports and energy drinks are 11gr, 7gr, 9gr and 16 gr per 100ml respectively. One gram of sugar is equivalent to 4 kcal or 16.7kJ (26).

#### Step 3 – Change in body weight, BMI & obesity status

##### Step 3a: Body weight

For children under 6, the change in weight for young children aged between 2 and 5 years was calculated based on the coefficients for change in weight per change in energy intake according to Long et al. (13). For boys, a change of 216 kJ/day would lead to 1 kilogram body weight change while for girls, one kilogram body weight change requires a change of 204 kJ/day (13). For children and adolescents aged 6–17 years, the equations from Hall et al. (27) were used. Hall and colleagues developed age- and gender-specific linear equations to predict the weight gain from a given energy imbalance. For children aged 6–17 years, the equations were kcal/day/kg = 68–2.5*age for males and kcal/day/kg = 62–2.2*age for females. For adults aged 18 and above, change in body mass was estimated using equations published by Swinburn et al. (28), which state that a daily increase in energy intake of 94 kJ/day is needed for a change in body weight of 1 kg in equilibrium for adults (93.0 kJ/day for men and 72.3 kJ/day for women). Generally, the above estimated weight change will take a year for a 50% achievement and three years for a 95% of total weight changes (28).

A Monte-Carlo simulation was applied at this stage to simulate the reduction of energy intake from SSBs to the average body weight change. In each simulation, the parameters are drawn from a normal distribution (28) with the final weight change computed by averaging the result from 10,000 iterations. The simulation was carried out separately by gender with the average value used in the following calculation in Step 3b. All the simulations were undertaken with Microsoft Excel macros developed in Visual Basic for Applications (VBA).

##### Step 3b: BMI index & obesity status

At this stage, the dataset of the National Survey on Non-communicable diseases risk factors (STEP) 2015 was used to develop a cohort simulation for further estimation for adult population aged 18 and above. Assuming there is no significant change in the height of all cohort participants, the BMI change was estimated based on the change in body weight after 3 years. The change in average BMI index was estimated for age and sex-specific groups of the STEP 2015 cohort and generalized to Vietnamese population aged 18-69. For children, the model was only able to estimate the projection of average weight change without further estimation to their BMI because children tend to experience rapid growth in their height which limits the validity of a BMI projection.

The prevalence of overweight and obesity was estimated using the threshold of Western Pacific Region of WHO for Asian population in which 23 ≤ BMI <25 is classified as overweight; 25≤BMI<30 as obesity grade I and BMI≥30 as obesity grade II (29). While other simulations were performed using Microsoft Excel macros, this step was carried out in STATA version 14.2 with the *svyset* command for complex survey data to extrapolate the estimation of overweight and obesity prevalence for Vietnamese population.

#### Step 4 - Change in burden of obesity-induced diseases

The findings in Asia Pacific Cohort Studies Collaboration 2005 (30) showed that for each 2 unit of BMI decrease, the risk of diabetes would reduce by 27% (95% confident interval: 23%-30%). The number of avoided diabetes cases was estimated using the relative reduction in diabetes prevalence. Probability of having diabetes with complications was applied on the model population as whole to estimate number of diabetes cases with and without complication that would be avoided after introduction of the SSBs tax, based on findings from Vietnamese health insurance claim database 2017 by Kiet Pham et al 2020 (31). The number of avoided cases were then multiplied with treatment cost for those diabetes cases with and without complications, taken from Kiet Pham et al 2020 (31). This outcome is limited for population aged 18 to 69 as the cohort simulation was conducted using STEP 2015 cohort of age 18-69. All data inputs & data sources can be found in Supplementary 1.

### Sensitivity Analysis

Probabilistic sensitivity analysis was performed using Monte Carlo simulation. The calorie-to-weight conversion factor was drawn using normal distribution, the relative risk of diabetes from BMI reduction is obtained from beta distribution. All simulations were carried out using the 95% confident interval and averaging from 10,000 iterations. The probabilistic analysis was later branched out into two models in which the Monte Carlo simulation employed two different sets of parameters. Model 1 performed the probabilistic analysis on weight change only while model 2 performed the extended probabilistic analysis on weight change and the risk reduction of diabetes because the risk reduction of diabetes based on BMI changes was highly sensitive to the calculation of diabetes’s burden. Additionally, one-way deterministic sensitivity analysis was carried out on the price elasticity at -1.0 and -0.8 (beside the price elasticity of -1.14 for base analysis). Findings of deterministic sensitivity analysis was included in Supplementary 2 for reference.

## MAIN FINDINGS

### 1. SSBs consumption & energy intake

**Table 2Error! Not a valid bookmark self-reference.** below shows five tax scenarios along with their impact on the SSB consumption. At the price elasticity of -1.14, the SSB consumption per capita per annum was estimated to reduce by 2.6 litres if the price increased by 5%; 5.1 litres if price increases by 11% and between 9.8-10.5 litres if price increase by 19-20%. Interestingly, although scenario 3 offered 19% price increase, the reduction in the amount of sugar consumed in this scenario (3.2 gr/ day/ person) would be higher than scenario 4 of 20% price increase (3.1 gr/ day/ person). This interesting result was mostly contributed from the reduction in high sugar content product lines (carbonates and sport drinks). Following the reduction of sugar consumed, scenario 3 continues to predict larger reduction in energy intake than scenario 4 and is almost of equivalent impact to scenario 5. Clearly, the three scenarios of 19-20% price increase were projected to offer much higher reduction in energy intake compared to scenario 1 (price increase of 5%) and scenario 2 (price increase of 11%). Further findings on price elasticity of -1.0 and - 0.8 could be found in Supplementary 2.

**Table 2:** Changes in SSBs’ consumption in five SSB tax scenarios.

| Tax designs | Scenario 1 | Scenario 2 | Scenario 3 | Scenario 4 | Scenario 5 |
| --- | --- | --- | --- | --- | --- |
|  | 10% ad-valorem | 3,500d/ litre | 60d/gr/100ml | 7,000d/ litre | 40% ad-valorem |
| Average price increase | 5% | 11% | 19% | 20% | 20% |
| Decrease in total SSB consumption (million litres/year) | 171.3 | 334.0 | 641.0 | 668.0 | 684.9 |
| Decrease in consumption (litre/person/ year) | 2.6 | 5.1 | 9.8 | 10.2 | 10.5 |
| Decrease in sugar consumed (gram/ day/ person) | 0.8 | 1.6 | 3.2 | 3.1 | 3.3 |
| Decrease in calorie intake from SSBs (kJ/ day/ person) | 13.7 | 26.2 | 54.0 | 52.4 | 54.9 |

### 2. Changes in body weight, BMI & obesity status

**Table 3** shows the projected body weight reduction, average BMI index and prevalence of overweight and obesity in different SSB tax scenarios. In scenarios of 20% price increase, the biggest predicted change in body weight was expected to reach about 0.57-0.59 kg for males and 0.73-0.76 kg reduced for females after 3 years of tax implementation. Noticeably, females could observe greater reduction of body weight change than males in all five tax scenarios. Corresponding to the change in body weight, the average BMI after SSB tax could vary up to 0.20 BMI unit between 5 scenarios (the variation would be 0.16 among men and 0.24 among women).

**Table 3:** Estimated weight changes and BMI index after 3 years of tax implementation.

| Tax designs | Scenario 1 | Scenario 2 | Scenario 3 | Scenario 4 | Scenario 5 |
| --- | --- | --- | --- | --- | --- |
|  | 10% *ad-valorem* | 3,500d/ litre | 60d/ gr/100ml | 7,000d/ litre | 40% *ad-valorem* |
| **Weight change (kg) - Mean (SD)** |  |  |  |  |  |
| *Children aged 2-17\** |  |  |  |  |  |
| Boys | 0.08 | 0.16 | 0.32 | 0.31 | 0.33 |
| Girls | 0.09 | 0.17 | 0.34 | 0.33 | 0.35 |
| *Adults aged 18+* |  |  |  |  |  |
| Male | 0.15 (0.01) | 0.28 (0.02) | 0.58 (0.03) | 0.57 (0.03) | 0.59 (0.03) |
| Female | 0.19 (0.01) | 0.36 (0.02) | 0.75 (0.04) | 0.73 (0.03) | 0.76 (0.04) |
| **Average BMI index after tax increase – Mean (SE)** |  |  |  |  |  |
| Both sexes | 21.89 (0.09) | 21.83 (0.09) | 21.69 (0.09) | 21.70 (0.09) | 21.69 (0.09) |
| Male | 21.92 (0.12) | 21.87 (0.12) | 21.76 (0.12) | 21.77 (0.12) | 21.76 (0.12) |
| Female | 21.86 (0.12) | 21.79 (0.12) | 21.62 (0.12) | 21.63 (0.12) | 21.62 (0.12) |
| **Projected overweight prevalence (23 ≤ BMI <25) – Mean (SE)** |  |  |  |  |  |
| *Baseline overweight prevalence: 17.9% (19.2% male; 16.5% female)* |  |  |  |  |  |
| Both sexes | 17.6%<br>(0.88%) | 17.0%<br>(0.84%) | 15.9%<br>(0.81%) | 15.9%<br>(0.81%) | 15.7%<br>(0.80%) |
|  | 10% ad-valorem | 3,500d/ litre | 60d/ gr/100ml | 7,000d/ litre | 40% ad-valorem |
| Male | 19.3%<br>(1.38%) | 18.6%<br>(1.33%) | 17.2%<br>(1.30%) | 17.2%<br>(1.30%) | 16.9%<br>(1.28%) |
| Female | 16.0%<br>(0.98%) | 15.4%<br>(0.98%) | 14.7%<br>(0.90%) | 14.7%<br>(0.90%) | 14.6%<br>(0.90%) |
| **Projected obesity grade I prevalence (25 ≤ BMI <30) – Mean (SE)** |  |  |  |  |  |
| *Baseline obesity grade I prevalence: 13.6% (13.1% male; 14.2% female)* |  |  |  |  |  |
| Both sexes | 13.2%<br>(0.81%) | 12.9%<br>(0.81%) | 12.4%<br>(0.81%) | 12.4%<br>(0.81%) | 12.4%<br>(0.81%) |
| Male | 12.6%<br>(1.19%) | 12.4%<br>(1.18%) | 12.2%<br>(1.18%) | 12.2%<br>(1.18%) | 12.2%<br>(1.18%) |
| Female | 13.8%<br>(1.02%) | 13.4%<br>(1.01%) | 12.6%<br>(0.97%) | 12.5%<br>(0.98%) | 12.6%<br>(0.97%) |
| **Projected obesity grade II prevalence (BMI ≥30) – Mean (SE)** |  |  |  |  |  |
| *Baseline obesity grade I prevalence: (3.4% (1.7% male; 5.1% female)* |  |  |  |  |  |
| Both sexes | 3.4%<br>(0.44%) | 3.3%<br>(0.43%) | 3.2%<br>(0.43%) | 3.2%<br>(0.43%) | 3.2%<br>(0.43%) |
| Male | 1.6%<br>(0.37%) | 1.6%<br>(0.37%) | 1.5%<br>(0.36%) | 1.5%<br>(0.36%) | 1.5%<br>(0.36%) |
| Female | 5.1%<br>(0.76%) | 5.0%<br>(0.74%) | 4.8%<br>(0.74%) | 4.9%<br>(0.74%) | 4.8%<br>(0.74%) |

\* No SD was provided for the reduction in weight among children as the result was estimated using cohort data, not individual-level data so the result cannot be extrapolated

**Table 3** also presents a lower figure of overweight and obesity compared to baseline prevalence. The smallest projected impact on the overweight and obesity prevalence was from scenario 1 at 17.6% (overweight), 13.2% (obesity grade I) and 3.4% (obesity grade II), respectively. The lowest projected prevalence was from scenario 3 and 5, estimated at 15.7%-15.9% (overweight), 12.4% (obesity grade I) and 3.2% (obesity grade II). Due to the small baseline figure, the projected absolute prevalence of obesity at BMI≥30 would not vary largely but stay from 2.8%-3.2% for both sexes, 1.1-1.5% for males, and 4.4-4.8% for females.

### 3. Changes in burden of obesity-induced diseases

**Table 4** presents results for the uncertainty analysis in which the 95% uncertainty intervals from the Monte Carlo estimation was reported in the parentheses. Overall, the SSB tax would reduce the average BMI of population from between 0.07 to 0.28 BMI units. The reduction of BMI among females was larger than males. This would give rise to a reduction of the diabetes prevalence by 0.03-0.12 percentage points compared to the base prevalence of 4.1%, which is equivalent to between approximately 20,000 – 80, 000 diabetes cases avoided. Females could have a bigger benefit compared to males due to a larger reduction in diabetes prevalence.

**Table 4:** Changes in the burden of diabetes mellitus induced by excess weight changes.

| Tax designs | Scenario 1 | Scenario 2 | Scenario 3 | Scenario 4 | Scenario 5 |
| --- | --- | --- | --- | --- | --- |
|  | 10% ad-valorem | 3,500d/ litre | 60d/ gr/100ml | 7,000d/ litre | 40% ad-valorem |
| **Average BMI reduction** |  |  |  |  |  |
| Both sexes | 0.07(0.0003) | 0.13 (0.0006) | 0.27 (0.0013) | 0.26 (0.0013) | 0.28 (0.0013) |
| Male | 0.06 (0.0002) | 0.11 (0.0003) | 0.22 (0.0007) | 0.22 (0.0006) | 0.23 (0.0007) |
| Female | 0.08 (0.0002) | 0.16 (0.0004) | 0.32 (0.0009) | 0.31 (0.0008) | 0.33 (0.0009) |
| **MODEL 1: Monte-Carlo simulation on calorie-to-weight conversion factor** |  |  |  |  |  |
| **Reduction in diabetes prevalence (percent point)** |  |  |  |  |  |
| Both sexes | 0.03% | 0.06% | 0.12% | 0.12% | 0.12% |
|  | (0.000%) | (0.000%) | (0.001%) | (0.001%) | (0.001%) |
| Male | 0.01% | 0.03% | 0.06% | 0.05% | 0.06% |
|  | (0.000%) | (0.000%) | (0.000%) | (0.000%) | (0.000%) |
|  | 10% ad-valorem | 3,500d/ litre | 60d/ gr/100ml | 7,000d/ litre | 40% ad-valorem |
| Female | 0.04% | 0.08% | 0.17% | 0.17% | 0.17% |
|  | (0.000%) | (0.000%) | (0.000%) | (0.000%) | (0.000%) |
| **No. of avoided diabetes case** | 20,270<br>(88) | 38,668<br>(175) | 79,705<br>(380) | 77,365<br>(380) | 80,992<br>(380) |
| **Healthcare cost saving (mil. USD)** | 6.94<br>(0.03) | 13.23<br>(0.06) | 27.28<br>(0.13) | 26.48<br>(0.13) | 27.72<br>(0.13) |
| **Healthcare cost saving (bil. VND)** | 155.19<br>(0.67) | 296.05<br>(1.34) | 610.25<br>(2.91) | 592.33<br>(2.91) | 620.10<br>(2.91) |
| **MODEL 2: Monte-Carlo simulation on calorie-to-weight conversion factor & diabetes risk reduction** |  |  |  |  |  |
| **Reduction in diabetes prevalence (percent point)** |  |  |  |  |  |
| Both sexes | 0.03% | 0.06% | 0.12% | 0.12% | 0.12% |
|  | (0.000%) | (0.000%) | (0.001%) | (0.001%) | (0.001%) |
| Male | 0.01% | 0.03% | 0.06% | 0.05% | 0.06% |
|  | (0.000%) | (0.001%) | (0.001%) | (0.001%) | (0.001%) |
| Female | 0.04% | 0.08% | 0.17% | 0.17% | 0.17% |
|  | (0.000%) | (0.001%) | (0.001%) | (0.001%) | (0.001%) |
| **No. of avoided diabetes case** | 20,386<br>(132) | 38,889<br>(253) | 80,161<br>(521) | 77,807<br>(505) | 81,455<br>(529) |
| **Healthcare cost saving (mil. USD)** | 6.98<br>(0.05) | 13.31<br>(0.09) | 27.44<br>(0.18) | 26.63<br>(0.17) | 27.88<br>(0.18) |
| **Healthcare cost saving (bil. VND)** | 156.08<br>(1.12) | 297.75<br>(2.01) | 613.73<br>(4.03) | 595.72<br>(3.80) | 623.64<br>(4.03) |

In model 1, the reduction in prevalence of diabetes corresponded to about 20,270 – 80,992 cases of diabetes avoided over the lifespan of modelled cohort. For both sexes, this could further save up to 155.2-620.1 billion VND (6.9 -27.2 million USD, 2021’s exchange) for the direct medical cost of diabetes treatment. Model 2 provided a similar estimate to model 1 for the change in diabetes burden. The number of avoided diabetes cases was slightly higher than model 1, ranging from 20,386 to 81,455 cases. The saving in health care cost estimated in model 2 is also greater, ranging from 156.1 – 623.6 billion VND (7.0-27.9 million USD, 2021’s exchange).

## DISCUSSION

Overall, the results indicated that projected SSB consumption levels reduced in relation to the application of higher tax rates. The Ministry of Finance-proposed option of 10% ad valorem (equivalent to a 5% price increase) was predicted to bring down the total SSB consumption by 171.3 million litres, equivalent to the reduction of 2.6 litres per capita per annum. And the reduction was expected to be four times higher in response to an average price increase of 19-20% in three highest tax plans (scenario 3 to 5). The reduction in total amount of consumption would be 9.8. 10.2 and 10.5 litres per capita per year in scenario 3, 4 and 5, respectively. Although scenario 3 with 19% price increase has less impact on the SSB consumption compared to scenario 4 & 5 of 20% price increase, the reduction in sugar intake from SSBs was almost equal at 3.1-3.3 gram/day/person in all three tax options. This could be explained by the effect of tax design on consumption behaviours including an industry response (through product reformulation) because the sugar-based specific tax has more power to reduce the consumption of high sugar-content products and promote the switch to lower sugar-content lines.

Reflecting the similar pattern of reduction in the amount of sugar consumed, the change in diet related NCD burden is predicted to show the most benefits when a sufficient level in price increase is reached. There is a clear difference in the prevalence of overweight and obesity grade I when applying a price increase from 5% to 11% and up to 19-20% (see

**Table 3** above). Although, the change in obesity grade II and type 2 diabetes were marginal, a noticeable gap in predicted reductions between different levels of price increase maintained (see

**Table 3** above). More importantly, at the same level of price increase at 19-20%, scenario 3 of sugar-based specific tax showed a higher benefit compared to the volume-based specific tax design (scenario 4) or almost equal to the ad valorem tax (scenario 5). This pointed to the fact that even at the same level of price increase, tax design targeted on high-sugar content product lines would have more health impacts. The design of specific tax based on sugar content or volume has recently proven successful at reducing consumption of high sugar-content SSB products and further encourage the switch to more healthy products (4). This finding should be highlighted when considering a new tax design for SSBs in Vietnam as well as other countries considering implementing SSB tax.

In comparison with previous simulation studies on SSB tax, the study results show consistent direction and compared to some countries, are close in magnitude of policy impacts on obesity status. At price increase of 20%, other simulation work on SSB tax reported the reduction in overweight and obesity prevalence varying from around 1% up to 3.8% such as in Thailand (15), Ireland (32), India (33), South African (17), Indonesia (18). This study estimated that at the same price increase, Vietnam would have about 1.2% drop in overweight and 0.2% reduction in obesity prevalence. The difference can be appreciable due to the different in country context of health trend and economic characteristics. A summary of previous simulation studies estimating SSB tax impacts on obesity prevalence can be found in Supplementary 3.

### Model limitations

Model limitations should not be ignored for further interpretation and implications. Firstly, the model outcomes are likely to offer a lower bound estimate of tax effects because only diabetes is modelled as a disease outcome and the body weight change was assumed from reduction in SSB consumption only, not accounting for reformulation of products to contain less sugar. Preliminary findings from Vietnam STEPS 2021 suggests that obesity burden is increasing rapidly compared to the 2015 figure (BMI≥25: 19.5% vs. 13.7%; diabetes: 7.5% vs. 4.1%) (34). Given the alarming trend of overweight and obesity and how long the policy formulation will take, our estimated impacts might only be able to slow down the increase rate of obesity rather than give actual reduction in overweight & obesity prevalence.

Additionally, model projection was also truncated within the cohort aged 18-69 so there may be substantially higher impacts when extended the modelled group to the whole population. Given the concern on childhood obesity in Vietnam during current time, further attempts on estimating SSB tax impact on childhood obesity explicitly is highly recommended. Additionally, one of the key parameters of calorie-to-weight conversion factor which originated from studies among Caucasian ethnicities might not provide a close approximation for Vietnamese population. Further, the assumption on the pass-on rate needs testing in the real world, but any reduction in this would likely have proportional impacts on health and cost consequences of an intervention.

Otherwise, this study presents another drawback that should be addressed in further studies if resources allow. The study did not provide estimates differentiated by potentially correlators such as socio-economic status (SES) or rural/urban residency. This is important not just in terms of estimating potential health effects where gradients related to SES may exist but also in terms of the distribution of the tax. International experiences showed that the acceptability of SSB taxes among both public and policymakers is greatly influenced by evidence related to the perceived equity of the tax burden (35, 36), and how the revenues will be used (35-37).

## CONCLUSION

The study’s findings are consistent with previous work, that suggest a tax on SSBs has the potential to reduce rates of over-weight/obesity and attain positive health effects as a result. By taxing SSBs, the benefit could be observed not only in the health of general population but also in the revenue gain and saving of healthcare cost for the government. The study focused on examining two determinants for developing SSB tax policy which are how and how much SSBs should be taxed.

Firstly, about the tax design, across three tiers of price increase, the impacts on consumption, health and economic would grow in line with the level of price increase. While 5% price increase would bring some benefits, increasing price up to 20% could bring a sizeable impact on the overweight and obesity rate. WHO recommends at least 20%, which is a good starting point for any country considering a tax (21, 38). Lawmakers and governmental authority must be aware that SSB tax must reach a sufficient level to obtain the expected benefits. Because the rapid economic growth can rapidly dominate the SSB tax increase, and potentially erase the deterrent effect of the tax on consumer behaviour. Moreover, when the price increase has been achieved a tax escalator should be incorporated into the tax if necessary to maintain its deterrent effect and adjusted regularly to eliminate the inflation impact.

Secondly, at the same price increase, the comparison of three tax designs has also shown some small differences in health benefits which the specific tax based on sugar density was predicted to be most effective on reducing sugar consumption from SSBs. The single tier based specific tax or ad valorem is familiar and straightforward for tax administration agency. The tax design based on sugar content is highly appraised for incentivizing industry to reformulation SSB products and have already been in practice in at least eight countries.

In conclusion, the study findings appear to offer strong support for the implementation of a SSB tax, which will slow down the increase rate of obesity burden and further reduce this burden if the policy is taken strongly. This is highly likely to benefit population health improvement through its contribution to a healthy food environment while generating considerable government revenue. The implementation of a SSB tax policy should be facilitated as multisector initiative (39, 40). There is a need to continue to develop and disseminate messages that promote healthy food choices (39, 40). Finally, it is important to keep in mind the study limitations, the nature of simulation work and noted uncertainties when considering the results and implications of the findings.

**Figure 1:**
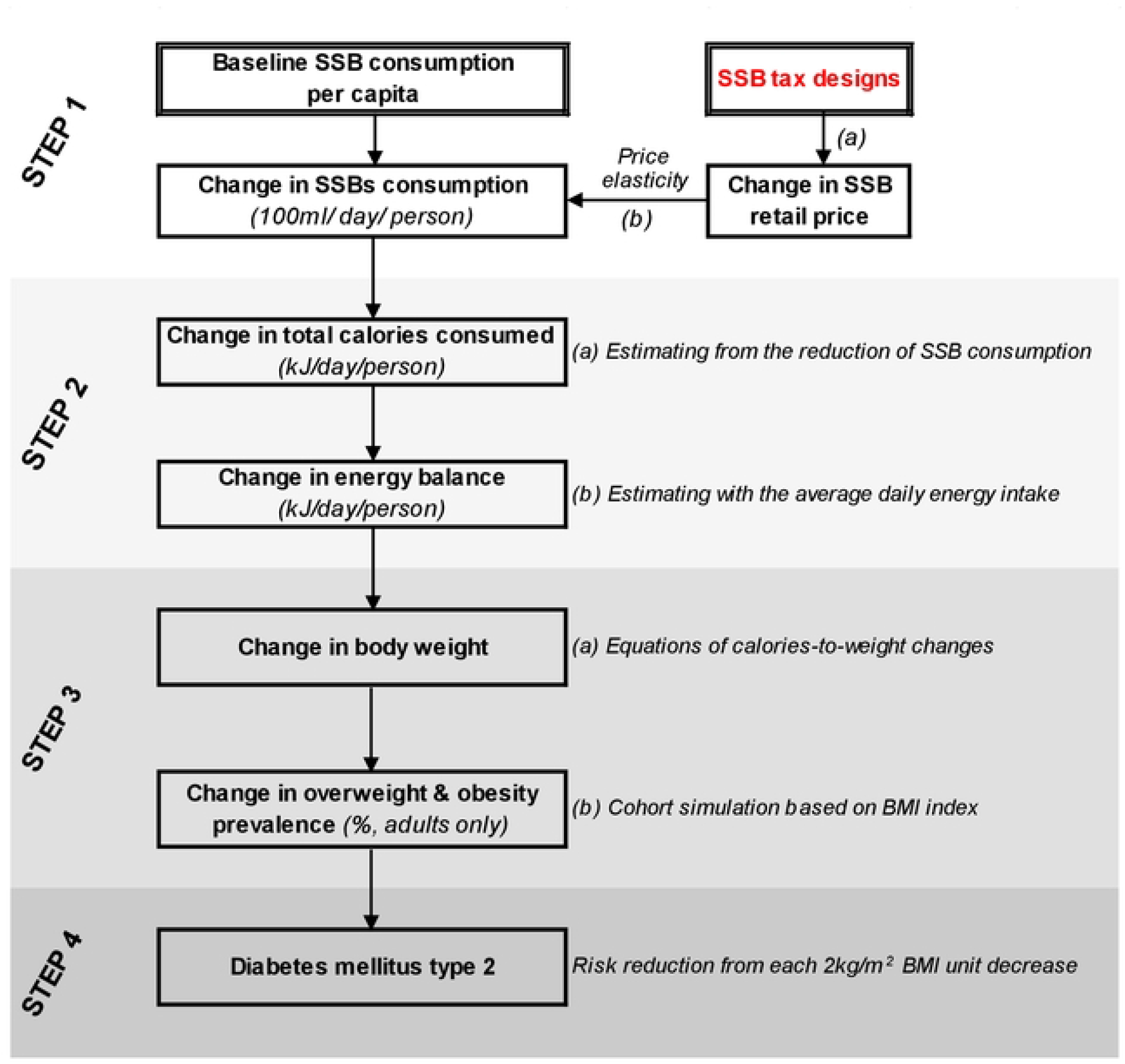
Model structure.

## Data Availability

All relevant data are within the manuscript and its Supporting Information files.

## Acknowledgement

The authors would like to express our thanks to Lam Tuan Nguyen from World Health Organization (Vietnam); Hoa Ngo, Fiona Watson, Courtney Peters & Jade Little from UNICEF; Doan Thu Huyen & Nguyen Van Huan from Global Health Advocacy Incubator for their valuable insights & comments throughout our work. A special thank goes to Professor Michael Donnelly (Queen’s University Belfast) for his time on reviewing and suggestions that has greatly improved this manuscript. The first author was funded by a PhD award from the School of Medicine, Dentistry and Biomedical Sciences, Queen’s University Belfast in memory of QUB Professors Dr Liam Murray and John Yarnell.

## SUPPLEMENTARY 1: Data inputs and assumptions

**Table S1** describes the main model inputs and assumptions with specific clarifications. Data for these inputs and assumption were drawn from the desk review and various available data sources as indicated.

**Table S1:** Model parameters & data sources.

| Indicators | Description | Data sources |
| --- | --- | --- |
| **Step 1: Change in SSB consumption** |  |  |
| Price elasticity | A parameter for estimating the change of SSB purchasing and consuming of when the price is increased from the taxation. | Linh Luong et al 2020 (23); literature review |
| Tax increase (%) | The relative change of SSB tax rate compared to the baseline rate to set up different tax scenarios | Shared data from the joint work of WHO & Health Bridge Canada |
| Baseline SSB consumption (litre) | Amount of SSB consumption in year 2020 separated by 4 categories (carbonates, juices, ready-to-drink tea/ coffee & sport drinks) | Global Data 2021 |
| SSB consumption per capita (100ml/ day/ person) | Amount of SSB consumption per capita in year 2020, separated by 4 categories (carbonates, juices, ready-to-drink tea/ coffee, energy & sport drinks) | Author's calculation for population age 2+ (using GSO Census 2019) |
| Population size | Population structure by age and sex, also representing the market size of SSB in Vietnam | GSO Census 2019 |
| **Step 2: Change in energy intake** |  |  |
| Sugar content | The average amount of sugar contained in each SSB lines, measured in gram/ 100ml | Paraje G. (WHO's expert report) (41) |
| Calorie density | The average calorie density in each SSB categories, measured in kJ/100ml | Author's calculation from sugar content |
| Baseline energy intake | The average daily intake of calories at baseline, measured as kJ/day/person | National Nutrition Survey 2019-2020 |
| **Step 3: Change in body weight, BMI & obesity status** |  |  |
| Conversion factors for calorie-to-weight | The corresponding change of weight due to the decreased intake of calorie consumed for children and adults | Literature review<br>(See more in Method section) |
| Baseline weight | The baseline weight of population by sex & age | STEP 2015 |
| Baseline height | The baseline height of population by sex & age | STEP 2015 |
| Baseline BMI | The baseline BMI of population by sex & age | STEP 2015 |
| BMI change | The absolute change between baseline BMI and the counterfactual BMI | Cohort simulation using STEP 2015 cohort |
| **Step 4: Change in burden of obesity-related diseases** |  |  |
| Risk reduction for diabetes | The relative reduction in risk of diabetes mellitus type 2 for each 2 kg/m<sup>2</sup> BMI unit decrease | Asia Pacific Cohort Studies Collaboration 2005 (30) |
| % of diabetes with complications | % of diabetes patients with complications | VSS 2017 (Kiet Pham et al 2020 (31)) |
| Medical cost for diabetes with complications | The average treatment cost of diabetes patients with complications, inflation adjusted to 2020 price (USD) | VSS 2017 (Kiet Pham et al 2020 (31)) |
| Medical cost for diabetes without complications | The average treatment cost of diabetes patients with complications, inflation adjusted to 2020 price (USD) | VSS 2017 (Kiet Pham et al 2020 (31)) |

## SUPPLEMENTARY 2: Sensitivity analysis result

**Table S2:** Deterministic sensitivity analysis on price elasticity at -1.0.

| Tax design |  | Scenario 1 | Scenario 2 | Scenario 3 | Scenario 4 | Scenario 5 |
| --- | --- | --- | --- | --- | --- | --- |
|  |  | 10% ad-valorem | 3,500d/ litre | 60d/ gr/100ml | 7,000d/ litre | 40% ad-valorem |
| **Weight change (kg)** |  |  |  |  |  |  |
| **Children aged 2-17** | Boys | 0.07 | 0.14 | 0.28 | 0.27 | 0.29 |
|  | Girls | 0.08 | 0.15 | 0.30 | 0.29 | 0.31 |
| **Adults aged 18+** | Male | 0.13 (0.01) | 0.25 (0.01) | 0.51 (0.03) | 0.50 (0.03) | 0.52 (0.03) |
|  | Female | 0.17 (0.01) | 0.32 (0.02) | 0.66 (0.03) | 0.64 (0.03) | 0.67 (0.03) |
| **Projected overweight prevalence (23 ≤ BMI <25) – Mean (SE)** |  |  |  |  |  |  |
| Both sexes |  | 17.7%<br>(0.88%) | 17.0%<br>(0.85%) | 16.5%<br>(0.83%) | 16.5%<br>(0.83%) | 16.4%<br>(0.82%) |
| Male |  | 19.5%<br>(1.37%) | 18.5%<br>(1.33%) | 18.0%<br>(1.32%) | 18.1%<br>(1.32%) | 17.8%<br>(1.32%) |
| Female |  | 15.9%<br>(0.98%) | 15.6%<br>(0.99%) | 15.0%<br>(0.95%) | 15.0%<br>(0.95%) | 14.9%<br>(0.95%) |
| **Projected obesity grade I prevalence (25 ≤ BMI <30) – Mean (SE)** |  |  |  |  |  |  |
| Both sexes |  | 13.2%<br>(0.81%) | 13.0%<br>(0.82%) | 12.7%<br>(0.82%) | 12.7%<br>(0.82%) | 12.7%<br>(0.82%) |
| Male |  | 12.6%<br>(1.19%) | 12.5%<br>(1.19%) | 12.4%<br>(1.18%) | 12.4%<br>(1.18%) | 12.4%<br>(1.18%) |
| Female |  | 13.8%<br>(1.02%) | 13.4%<br>(1.01%) | 13.1%<br>(1.01%) | 13.1%<br>(1.01%) | 13.1%<br>(1.01%) |
| **Projected obesity grade II prevalence (BMI ≥30) – Mean (SE)** |  |  |  |  |  |  |
| Both sexes |  | 3.4%<br>(0.44%) | 3.3%<br>(0.43%) | 3.3%<br>(0.43%) | 3.3%<br>(0.43%) | 3.3%<br>(0.43%) |
| Male |  | 1.6%<br>(0.37%) | 1.6%<br>(0.37%) | 1.6%<br>(0.37%) | 1.6%<br>(0.37%) | 1.6%<br>(0.37%) |
| Female |  | 5.1%<br>(0.76%) | 5.0%<br>(0.74%) | 4.9%<br>(0.74%) | 4.9%<br>(0.74%) | 4.9%<br>(0.74%) |
| **% reduction in diabetes prevalence (percentage point)** |  |  |  |  |  |  |
| Both sexes |  | 0.03%<br>(0.000%) | 0.05%<br>(0.000%) | 0.09%<br>(0.001%) | 0.08%<br>(0.001%) | 0.09%<br>(0.001%) |
| Male |  | 0.01%<br>(0.000%) | 0.02%<br>(0.000%) | 0.04%<br>(0.001%) | 0.04%<br>(0.001%) | 0.04%<br>(0.001%) |
| Female |  | 0.04%<br>(0.000%) | 0.07%<br>(0.001%) | 0.12%<br>(0.001%) | 0.12%<br>(0.001%) | 0.12%<br>(0.001%) |
| **Number of avoided diabetes case** |  | 17,804<br>(88) | 34,023<br>(176) | 56,080<br>(264) | 54,437<br>(264) | 56,989<br>(264) |
| **Healthcare cost saving (mil. USD)** |  | 6.11<br>(0.04) | 11.68<br>(0.07) | 19.25<br>(0.12) | 18.69<br>(0.12) | 19.56<br>(0.12) |
| **Healthcare cost saving (bil. VND)** |  | 136.71<br>(0.89) | 261.26<br>(1.57) | 430.64<br>(2.68) | 418.02<br>(2.68) | 437.62<br>(2.68) |

**Table S3:**
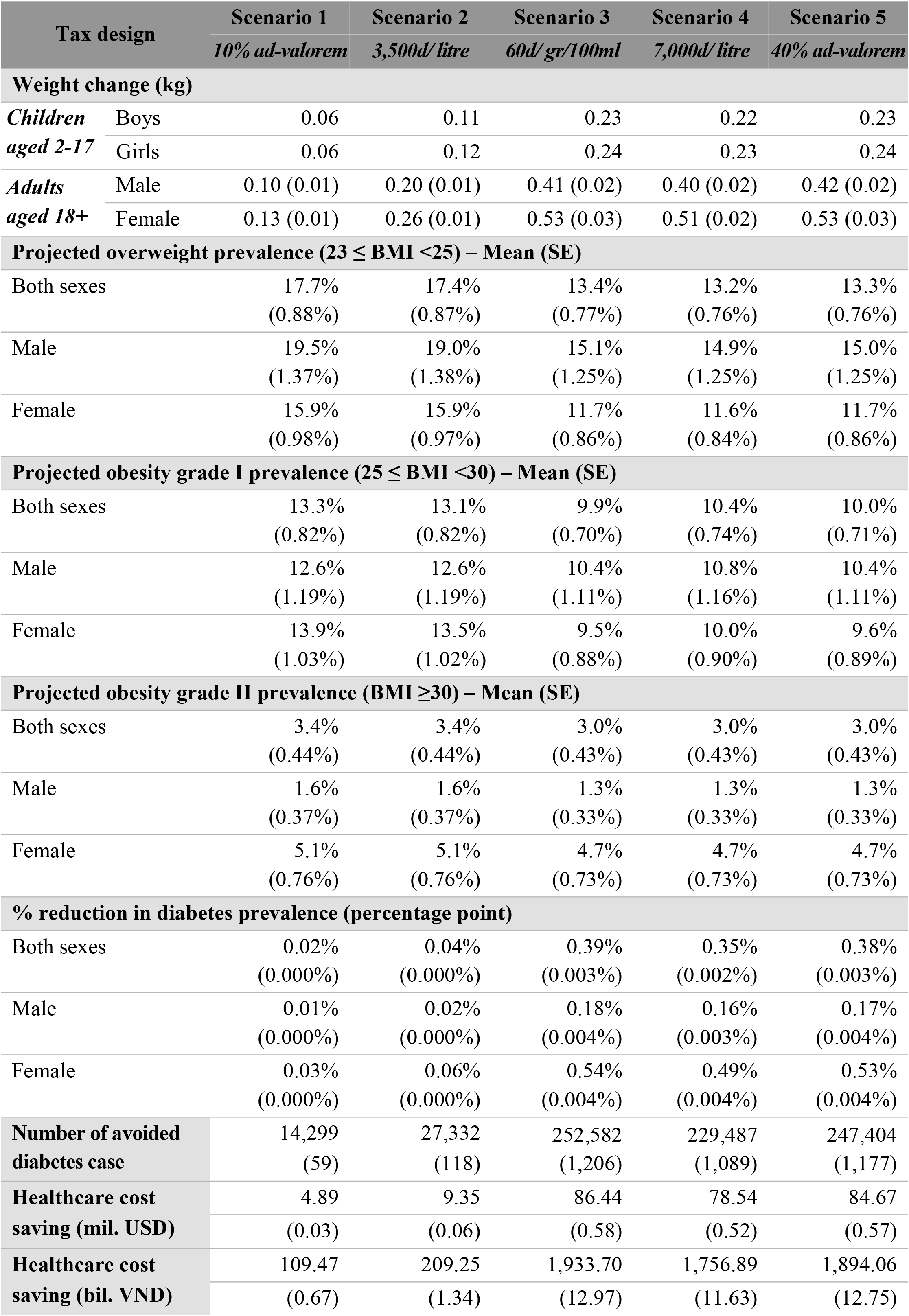
Deterministic sensitivity analysis on price elasticity at -0.8.

| Tax design |  | Scenario 1 | Scenario 2 | Scenario 3 | Scenario 4 | Scenario 5 |
| --- | --- | --- | --- | --- | --- | --- |
|  |  | 10% ad-valorem | 3,500d/ litre | 60d/ gr/100ml | 7,000d/ litre | 40% ad-valorem |
| **Weight change (kg)** |  |  |  |  |  |  |
| **Children aged 2-17** | Boys | 0.06 | 0.11 | 0.23 | 0.22 | 0.23 |
|  | Girls | 0.06 | 0.12 | 0.24 | 0.23 | 0.24 |
| **Adults aged 18+** | Male | 0.10 (0.01) | 0.20 (0.01) | 0.41 (0.02) | 0.40 (0.02) | 0.42 (0.02) |
|  | Female | 0.13 (0.01) | 0.26 (0.01) | 0.53 (0.03) | 0.51 (0.02) | 0.53 (0.03) |
| **Projected overweight prevalence (23 ≤ BMI <25) – Mean (SE)** |  |  |  |  |  |  |
| Both sexes |  | 17.7%<br>(0.88%) | 17.4%<br>(0.87%) | 13.4%<br>(0.77%) | 13.2%<br>(0.76%) | 13.3%<br>(0.76%) |
| Male |  | 19.5%<br>(1.37%) | 19.0%<br>(1.38%) | 15.1%<br>(1.25%) | 14.9%<br>(1.25%) | 15.0%<br>(1.25%) |
| Female |  | 15.9%<br>(0.98%) | 15.9%<br>(0.97%) | 11.7%<br>(0.86%) | 11.6%<br>(0.84%) | 11.7%<br>(0.86%) |
| **Projected obesity grade I prevalence (25 ≤ BMI <30) – Mean (SE)** |  |  |  |  |  |  |
| Both sexes |  | 13.3%<br>(0.82%) | 13.1%<br>(0.82%) | 9.9%<br>(0.70%) | 10.4%<br>(0.74%) | 10.0%<br>(0.71%) |
| Male |  | 12.6%<br>(1.19%) | 12.6%<br>(1.19%) | 10.4%<br>(1.11%) | 10.8%<br>(1.16%) | 10.4%<br>(1.11%) |
| Female |  | 13.9%<br>(1.03%) | 13.5%<br>(1.02%) | 9.5%<br>(0.88%) | 10.0%<br>(0.90%) | 9.6%<br>(0.89%) |
| **Projected obesity grade II prevalence (BMI ≥30) – Mean (SE)** |  |  |  |  |  |  |
| Both sexes |  | 3.4%<br>(0.44%) | 3.4%<br>(0.44%) | 3.0%<br>(0.43%) | 3.0%<br>(0.43%) | 3.0%<br>(0.43%) |
| Male |  | 1.6%<br>(0.37%) | 1.6%<br>(0.37%) | 1.3%<br>(0.33%) | 1.3%<br>(0.33%) | 1.3%<br>(0.33%) |
| Female |  | 5.1%<br>(0.76%) | 5.1%<br>(0.76%) | 4.7%<br>(0.73%) | 4.7%<br>(0.73%) | 4.7%<br>(0.73%) |
| **% reduction in diabetes prevalence (percentage point)** |  |  |  |  |  |  |
| Both sexes |  | 0.02%<br>(0.000%) | 0.04%<br>(0.000%) | 0.39%<br>(0.003%) | 0.35%<br>(0.002%) | 0.38%<br>(0.003%) |
| Male |  | 0.01%<br>(0.000%) | 0.02%<br>(0.000%) | 0.18%<br>(0.004%) | 0.16%<br>(0.003%) | 0.17%<br>(0.004%) |
| Female |  | 0.03%<br>(0.000%) | 0.06%<br>(0.000%) | 0.54%<br>(0.004%) | 0.49%<br>(0.004%) | 0.53%<br>(0.004%) |
| **Number of avoided diabetes case** |  | 14,299<br>(59) | 27,332<br>(118) | 252,582<br>(1,206) | 229,487<br>(1,089) | 247,404<br>(1,177) |
| **Healthcare cost saving (mil. USD)** |  | 4.89<br>(0.03) | 9.35<br>(0.06) | 86.44<br>(0.58) | 78.54<br>(0.52) | 84.67<br>(0.57) |
| **Healthcare cost saving (bil. VND)** |  | 109.47<br>(0.67) | 209.25<br>(1.34) | 1,933.70<br>(12.97) | 1,756.89<br>(11.63) | 1,894.06<br>(12.75) |

## SUPPLEMENTARY 3: Comparison of current findings with previous simulation works

**Table S4:** Potential health impacts of SSB tax from previous simulation works.

| Country | Model characteristics | Tax scenario | Health benefits |
| --- | --- | --- | --- |
| Current study | Micro, static; Adult 18-69; prediction horizon of 1-3 years | Tax increase: 60d/gr/100ml<br>Price increase: 19%<br>PE = -1.14 | • Overweight *: ↓1.2%;<br>• Obesity *: ↓0.20%;<br>• Diabetes: ↓0.12% |
| Ireland (2013) (32) | Macro, static; Adults 18-75+; prediction horizon of 3 years | Tax increase: 10%<br>Price increase: 9%<br>PE = -0.9 | • Obesity: ↓1.3%<br>Overweight: ↓0.7% |
| India (2014) (33) | discrete micro simulation: hypothetical adult population aged 25-65; prediction horizon of 10 years | Tax increase: 20%<br>Price increase: 20%<br>PE = -0.94 | • Overweight & obesity: ↓3.0% (1.6-5.9);<br>• Diabetes: ↓1.6% (1.2-1.9) |
| South African (2014) (17) | Macro, static; Adults aged 15+; prediction horizon of 3 years | Tax increase: 20%<br>Price increase: 20%<br>PE = -1.299 | • Obesity prevalence: ↓3.8% (males); ↓2.4% (females) |
| Indonesia (2018) (18) | Macro, multistate life table based Markov model; Population 0-95+ years old; prediction horizon of a lifetime | Tax increase: approx. 20%<br>Price increase: approx. 20%<br>PE = -1.13 to -1.34 | • Overweight: ↓2.9% (males); ↓1.4% (females)<br>• Obesity: ↓7.3% (males), ↓3.9% (females); |
| Zambia (2020) (42) | Macro lifetable based Markov model; Adults aged 15-65+; prediction horizon of 40 years | Tax increase: 25%<br>Price increase: 25%<br>PE = -1.30 | • Obesity prevalence: ↓0.49% (0.41-0.57) |
| Thailand (2021) (15) | Macro, static; Children 3-17 & adults 18+; prediction horizon of 1-3 years | Tax increase: 11%, 20%, 25%<br>Price increase: 11%, 20%, 25%<br>PE = 1.30 | • Obesity prevalence: ↓1.73%, ↓3.83%, and ↓4.91%, respectively |

\* To be consistent with other studies, the study results on overweight are reported as of $25 \leq BMI < 30$ and obesity as of $BMI \geq 30$ . ↓ indicates the reduction on prevalence

